# Contextual factors shaping the delivery of routine immunisation services in the Mumbunda Health Zone, Democratic Republic of the Congo: a multiple case study of head nurses’ experiences

**DOI:** 10.64898/2026.09.16.26363244

**Authors:** Chadrack Kabeya Diyoka, Angèle Musau Nkola, Angelique Salima Saidi, Pascal Pambi Mukanga, Pierre Cisuaka Nkongolo, Diane Muantama Balimo, Hendrick Mbutshu Lukuke, Didier Chuy Kalombola

## Abstract

**Objectives:** To analyse how contextual factors shape the delivery of routine Expanded Programme on Immunisation (EPI) services in the urban Mumbunda Health Zone, Democratic Republic of the Congo, from the perspective of head nurses.

**Design:** Multiple case study conducted from May to June 2026.

**Setting:** Seven health areas in the urban Mumbunda Health Zone, Democratic Republic of the Congo.

**Participants:** Seven head nurses selected purposively; four participants took part in additional follow-up interviews.

**Methods:** Semi-structured individual interviews were analysed using reflexive thematic analysis, with systematic within-case and cross-case comparisons. Findings were organised using the four contextual dimensions of Leichter’s framework.

**Results:** Nine subthemes and one cross-cutting theme were identified. Situational factors comprised population mobility, seasonality and time constraints that affected access to, and monitoring of, immunisation. Structural factors included peripheral governance, microplanning and information flows; functional staff availability; operational and logistical resources; and community organisation. Cultural factors included professional norms, variable uptake of EPI requirements, perceptions of vaccination and language-related influences on trust. Exogenous factors reflected dependence on decisions at health-zone level and on external resources, alongside the transfer of some routine costs to facilities and health workers. Across all four dimensions, participants described compensatory local adaptations that helped sustain service delivery despite these constraints.

**Conclusions:** Strengthening routine immunisation requires participatory microplanning, predictable financing for routine operational costs and context-sensitive support for facility teams and community health workers. Policy should enable useful local adaptation while reducing reliance on health workers’ personal resources to sustain day-to-day service delivery.

**Strengths and limitations of this study:**

- The multiple case design enabled systematic examination of similarities and differences within and across seven health areas in the same urban health zone.
- Follow-up interviews with four participants were used to clarify, elaborate and refine their accounts.
- Leichter’s framework provided an explicit structure for analysing contextual factors while allowing a cross-cutting theme to emerge inductively.
- The findings reflect the perspectives of seven head nurses and do not directly represent those of other EPI stakeholders.
- Data were collected in a single health zone over a limited period, restricting examination of variation across settings and over time.

## Introduction

Globally, the Expanded Programme on Immunisation (EPI), launched by the World Health Organisation (WHO) in 1974, is among the public health interventions that have contributed most substantially to child survival (1,2). Modelling of its first 50 years estimated that vaccination averted approximately 154 million deaths between 1974 and 2024, the vast majority among children younger than 5 years (3). Despite these gains, millions of children remain unvaccinated or under-vaccinated, and progress remains uneven both between and within countries. These disparities suggest that vaccine availability and standardised EPI strategies alone are insufficient to ensure effective, continuous and equitable delivery of immunisation services.

How these strategies are implemented depends substantially on the context in which they are embedded (4–6). Context is therefore not merely an external backdrop to the EPI; it shapes actors’ scope for action, resource mobilisation, relationships across levels of the health system and adaptations made during implementation (4–6). Leichter’s (1979) framework provides one way of examining this complexity by distinguishing situational, structural, cultural and exogenous factors that shape the implementation of policies and interventions (7).

International experience shows that immunisation programmes operating under broadly similar guidelines can achieve different results according to their implementation context. In a multiple case study in Nepal, Senegal and Zambia, Sakas et al. (2023) found that high routine immunisation performance was associated with a combination of effective governance, supportive political environments, health-system strengthening, community engagement and adaptation of EPI activities to local realities (8). In Pakistan, Haq et al. (2019) showed that EPI challenges arose from interactions among fixed and outreach delivery strategies, human resources, the availability of reliable demographic data, coordination, financing, polio-eradication priorities and the sociopolitical environment (9). More recently, Lawrence et al. (2025) identified contextual differences in Malawi and Mozambique relating to team composition, vaccine availability, vaccination-card policies and the organisation of outreach activities, showing that apparently similar barriers may operate differently across local service configurations (10). Evidence from low- and middle-income countries therefore supports a multidimensional view in which health-system, geographical, socioeconomic and social factors interact rather than operate in isolation.

This perspective is particularly relevant in the WHO African Region. District-level studies in Ethiopia, Cameroon and Ghana showed that differences in immunisation performance were not explained solely by infrastructure or basic resource availability. Districts that improved coverage were more clearly distinguished by management practices, use of data, implementation strategies, community engagement and the capacity to adapt services to local needs (11). Similarly, a recent review of determinants of vaccination implementation in the African Region found contextual factors operating across multiple system levels through complex interdependencies (12). The regional literature therefore highlights substantial subnational heterogeneity, with determinants and their effects varying across regions, urban and rural settings and population groups, thereby limiting the appropriateness of uniform approaches.

This challenge is particularly marked in the Democratic Republic of the Congo (DRC). According to the 2023–2024 DRC Demographic and Health Survey, only 21% of children aged 12–23 months had received all basic antigens, while 23% had received no vaccination (13). These low levels coexist with substantial provincial and intraprovincial disparities. Evidence from the DRC also indicates that the problem extends beyond household demand. The Mashako Plan identified weaknesses in coordination, service delivery, vaccine availability, monitoring and data use; before its implementation, 36% of the health areas assessed conducted fewer than one vaccination session per week and 92% had experienced at least one stock-out (14). Le Gargasson et al. (2014) also documented constraints in financing, budgetary processes, disbursement and the flow of resources to operational levels (15). Other studies have identified inequalities related to poverty, education, distance, quality of reception, reminders and service organisation (16–18). Routine EPI delivery in the DRC therefore reflects the interaction of political, organisational, financial, logistical, social and territorial factors whose effects vary across local contexts.

The Mumbunda Health Zone in Lubumbashi illustrates the limitations of relying on aggregate indicators alone. In 2025, reported overall administrative immunisation coverage reached 111%, above the EPI target of 95%, while concealing substantial variation between antigens (19). A previous study in this health zone identified several bottlenecks affecting routine immunisation, particularly in governance, financing, resource availability and service delivery, as well as their underlying causes (20). Identifying bottlenecks, however, primarily indicates what is not functioning; it provides less insight into the contextual conditions through which these bottlenecks arise and assume different forms across health areas. This distinction is important where the same EPI guidelines are implemented in operational units facing different organisational, territorial and social realities.

These processes remain insufficiently documented in the Congolese literature. Existing studies provide comparatively strong evidence on individual determinants of vaccination, zero-dose and under-vaccinated children, geographical inequalities and specific health-system constraints, but less evidence on the operational mechanisms through which these conditions combine at facility level.

Within the Congolese health system, head nurses responsible for health areas occupy a pivotal position at the interface between the Health Zone Central Office, health facilities, service providers and communities (21). Their experience can illuminate not only the presence of contextual factors but also their interactions, their practical manifestations in service delivery and the adaptations developed in response.

This study therefore aimed to analyse how contextual factors shape the delivery of routine EPI services across health areas in the Mumbunda Health Zone, from the perspective of head nurses, using the situational, structural, cultural and exogenous dimensions of Leichter’s framework (1979).

## Methods

### Study design and period

We conducted a qualitative multiple case study. Data collection for the initial phase took place from May to June 2026. Each selected health area constituted a case through which the context of routine EPI implementation was examined.

### Study setting

The study was conducted in the urban Mumbunda Health Zone in Lubumbashi, Haut-Katanga Province, Democratic Republic of the Congo. The health zone falls under the Haut-Katanga Provincial Health Division and was created in 2006 following subdivision of the former Lubumbashi Health Zone. It covers an estimated area of 72 km² (19,22) (figure 1).

**Figure 1.**
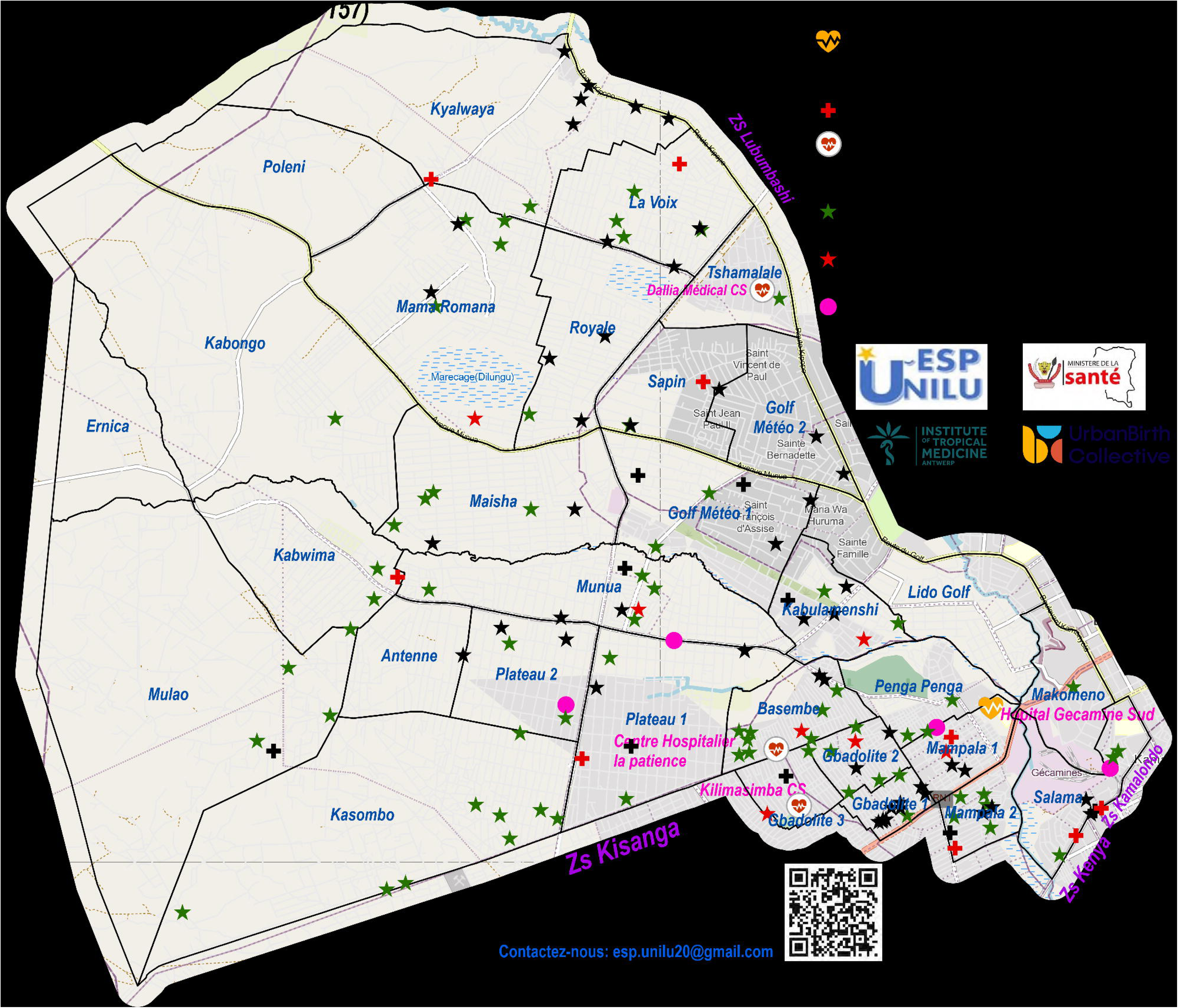
Map of the Mumbunda Health Zone.

The population of the Mumbunda Health Zone was projected to increase from 568,357 inhabitants in 2025 to 624,467 in 2026. Children younger than 5 years represented approximately 4% of the population (about 24,979 children), forming an important target population for maternal and child health interventions. The health zone comprises 30 health areas and 186 health facilities; 142 facilities are integrated into the national DHIS2 health information system and 98 appear on the official health-zone list. The facilities include community health centres, state-run health centres, private facilities and faith-based facilities. Their geographical distribution is uneven, with greater concentration in densely populated urban sectors and relatively lower coverage in some peripheral areas (19).

### Selection of health areas and cases

In the DRC, the health zone constitutes the operational level of the health system and is subdivided into health areas coordinated by a health-zone management team (23). Each health area is organised around a health centre that delivers the minimum package of services to its catchment population (23–25). In urban settings, this organisation coexists with numerous public, private and faith-based facilities, making coordination among the main health centre, satellite facilities and the community particularly important (26). In each selected health area, the main health centre and one satellite facility were considered. The main health centre, led by the head nurse, coordinates the health area and acts as an interface between the Health Zone Central Office, satellite facilities and the community. For the quantitative component of the broader study, the sampling frame comprised 98 operational health facilities registered in DHIS2 and included on the official health-zone list. A sample of approximately 30% of these facilities (about 29 facilities) was selected using the approach applied in the initial study (27). Fifteen health areas were then selected to represent four profiles combining access to and use of immunisation services: good access– good use, good access–poor use, poor access–good use and poor access–poor use.

### Conceptual and analytical framework

The broader research programme adopted a systems perspective in which vaccination coverage was treated as an outcome of service delivery within the local health system. Service delivery was understood to depend on the coordination of stewardship, financing and human, material, logistical and information resources, all operating within the values and principles of the health system and a defined context (28–30). In the first study, this model structured a quantitative assessment of EPI functions and identified major bottlenecks in vaccination service delivery (20). Those findings informed the topics and issues explored in greater depth in the qualitative component reported here.

For the present analysis, we used Leichter’s conceptual framework to organise and interpret contextual influences (7,31). The framework distinguishes four dimensions of context: (i) situational factors, including events or circumstances that may disrupt implementation; (ii) structural factors, including system organisation and resources; (iii) cultural factors, including beliefs, norms and practices; and (iv) exogenous factors, including directives, decisions and support originating from higher levels or external partners. Together, these factors shape the conditions under which local health-system resources, governance, financing, information and actors are mobilised, thereby influencing delivery of routine EPI services and, ultimately, vaccination coverage.

### Participants, sampling and data collection

The study population comprised head nurses responsible for health areas involved in routine EPI delivery. Participants were selected purposively. Sampling proceeded iteratively across cases and was guided by analytical saturation, resulting in seven participants. Eligibility required current responsibility for a selected health area and provision of written informed consent.

Data consisted of participants’ accounts and perspectives obtained through semi-structured individual interviews lasting approximately 40 minutes to 2 hours. The principal investigator conducted all interviews face to face using an interview guide after obtaining written informed consent. Fifteen head nurses from the selected health areas were invited to participate in the qualitative phase of the broader study and all agreed; interviews for the present analysis continued until analytical saturation was reached (n=7) (Table 1). Four participants (IT1, IT4, IT5 and IT6) subsequently took part in two rounds of follow-up interviews, conducted from 22 to 28 May 2026 and from 18 to 21 June 2026, to clarify, elaborate or verify points arising during analysis (32). Interviews were audio-recorded using an Android smartphone recorder. Analytical saturation was considered reached when additional coding and analysis no longer generated substantive new codes, themes, patterns or relationships likely to alter interpretation of the phenomenon under study (33). Consistent with Naeem et al. (2024), subsequent data were used primarily to confirm, enrich or refine the themes and relationships already identified (33).

**Table 1.** Characteristics of participants.

| Participant characteristic | n |
| --- | --- |
| Individual interviews with head nurses | 7 |
| Total participants in the qualitative study | 7 |
| Head nurses | 7 |

### Data analysis

Audio recordings were listened to repeatedly and transcribed verbatim in Microsoft Word. Interviews conducted in Swahili were translated into French during transcription. Data were analysed manually using reflexive thematic analysis (34,35), with systematic comparison of similarities and differences within and across cases. Credibility, dependability, transferability and confirmability were considered throughout data collection and analysis. Microsoft Word and Excel 2019 and VLC media player 3.0.21 were used to support data management and analysis.

### Ethical considerations

In accordance with institutional procedures for specialisation studies at the School of Public Health, University of Lubumbashi, the study was deemed exempt from review by a research ethics committee. The School of Public Health authorised the study through a research permit issued on 3 November 2025 (Appendix 1), intended to ensure scientific and ethical compliance of the protocol. The Mumbunda Health Zone Central Office and participating health facilities were subsequently informed about the study objectives and data-collection procedures. Participation was voluntary and written informed consent was obtained from all participants.

The study was conducted in accordance with the ethical principles of the Declaration of Helsinki. Participants were informed of their right to decline participation or withdraw at any time without consequence. Recordings, transcripts and field notes were transferred from the recording devices to password-protected storage on a computer and an external hard drive, after which the source copies were deleted. During transcription, data were anonymised or pseudonymised and direct identifiers were removed. Before anonymisation, participants were given the opportunity to review their statements and request correction or deletion. Data were stored and managed securely to protect confidentiality.

## Results

### Participant characteristics

Seven head nurses participated; five were women and two were men. Participants were aged 34–55 years, with a median age of 45 years (IQR 40–50). Their length of service ranged from 10 to 27 years, with a median of 19 years (IQR 17.5–22.5) (table 2).

**Table 2.** Participant characteristics.

|  |  |
| --- | --- |
| Sex | n=7 |
| Female | 5 |
| Male | 2 |
| Length of service, median (IQR), years | 19 (17.5-22.5) |
| Age, median (IQR), years | 45 (40-50) |

Contextual factors shaping routine EPI service delivery in the Mumbunda Health Zone

Using Leichter’s four contextual dimensions as the initial analytical structure, nine subthemes and one cross-cutting theme were identified (table 3 and figure 2).

**Figure 2.**
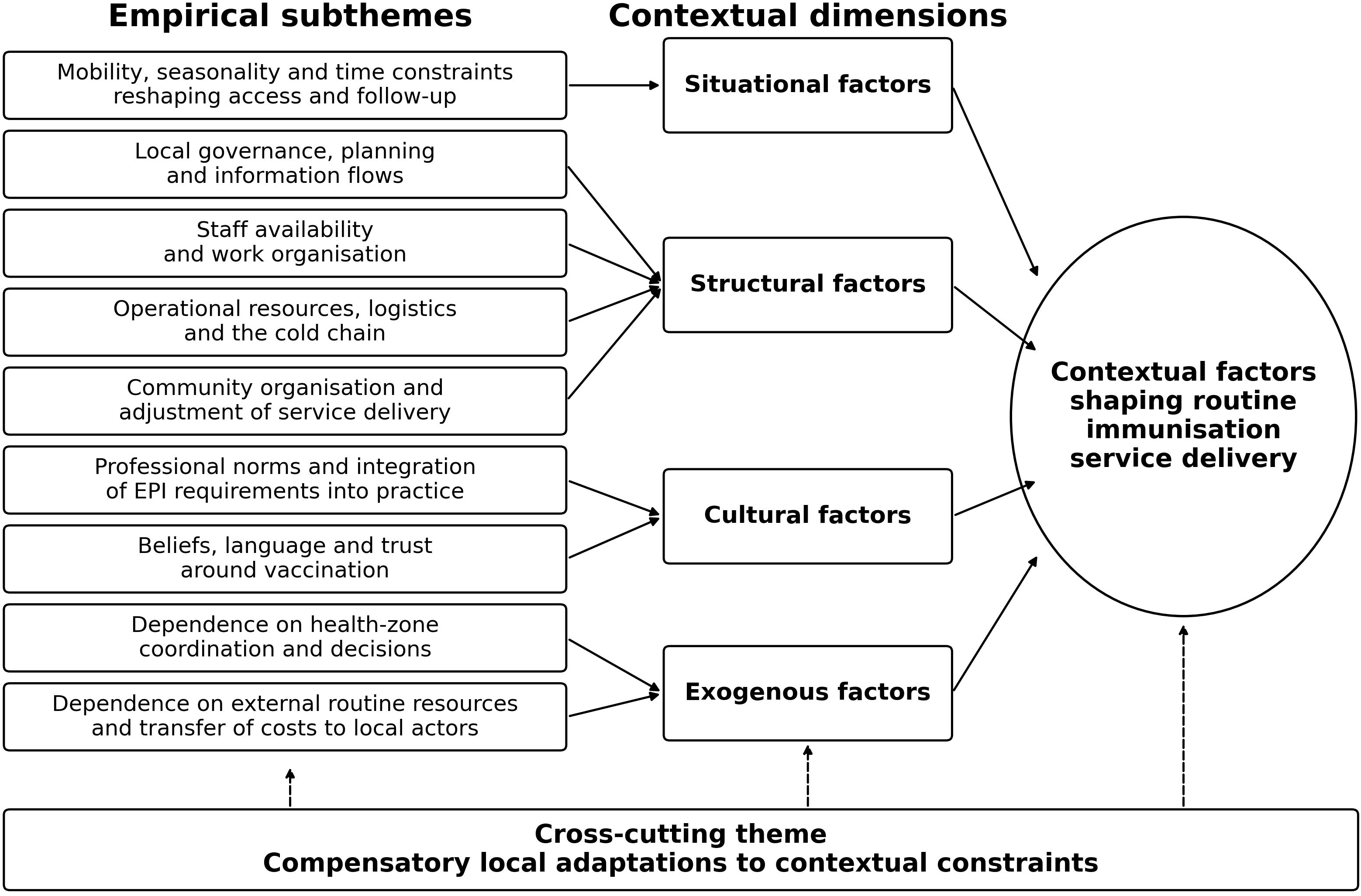
Simplified coding tree.

**Table 3.** Leichter contextual dimensions and corresponding empirical subthemes.

| Leichter contextual dimension | Empirical subtheme |
| --- | --- |
| Situational | Mobility, seasonality and time constraints reconfiguring access and monitoring |
| Structural | Peripheral governance, planning and information flow |
|  | Functional staff availability and work organisation |
|  | Operational resources, logistics and the cold chain |
|  | Organisation of the community interface and adaptation of service delivery |
| Cultural | Professional norms and variable uptake of EPI requirements |
|  | Perceptions, language and trust surrounding vaccination |
| Exogenous | Vertical dependence on coordination and decisions at health-zone level |
|  | External dependence on routine resources and transfer of costs to the periphery |
| Cross-cutting | Compensatory local adaptations to contextual constraints |

### Theme 1: Situational factors

One subtheme was identified: mobility, seasonality and time constraints reconfiguring access and monitoring.

### Subtheme 1: Mobility, seasonality and time constraints reconfiguring access and monitoring

Population movement across health-area boundaries meant that administrative catchment areas did not always correspond to families’ actual mobility patterns. Children from other health areas or health zones were regularly seen in some facilities, complicating the monitoring of cohorts assigned to each health area. Mobility was only one component of this territorial instability: seasonal conditions could also substantially alter physical access to some sites.

> “I also receive children from Kabulasmeshi, from Kisanga, outside the health area. […] There are others who come here to give birth just to get the BCG vaccine. They go elsewhere, to other facilities.” HN 5 “I take the registers, I check the addresses. At one point, I realized that about 40% come from outside the health area. […] There are those who come from outside the health areas. There are even those who come from outside the health zones.” HN 4 “Here in our neighborhood, we have many children who haven’t received any doses and are under-vaccinated. Many people move here […] when they arrive, they bring their children for a consultation; when you ask about their vaccination status, you learn that the child isn’t vaccinated. […] Here, there’s also the problem of accessibility during the rainy season. We can’t get through… The facility is in the Potager village; If it rains, you should know that you won’t be able to vaccinate, at least not those outside the health zone or the health area. […] Some people, because there’s flooding here. Even we can’t go out. HN 7 “We have our vaccination schedule, others call to say they’ll be late. […] If you’re not too busy, you can help them; otherwise, we’ll send them to the next session. […] If I’m alone at the station, I’ve already vaccinated, and I have a case of childbirth, I’ll prioritise that person, and she has to wait.” HN 6

### Theme 2: Structural factors

Four subthemes were identified: peripheral governance, planning and information flow; functional staff availability and work organisation; operational resources, logistics and the cold chain; and organisation of the community interface and adjustment of service delivery.

### Subtheme 2: Peripheral governance, planning and information flow

Participants described several governance mechanisms relevant to EPI delivery, including microplanning, monitoring, data validation, meetings and supervision; however, their translation into routine practice varied across health areas.

> “Yes, every month, at the end of each month, I go to my data-validation meetings. […] I hold monitoring meetings with the nurses in charge of the different facilities to discuss the data. […] If we see that the data is very low compared to our population, we ask the RECO, or ourselves, to raise awareness among mothers […] to come […] to have their children vaccinated.” HN 2 “At my place, the microplan is only available at the main facility; we’ve never taken the initiative to distribute it, to give others the opportunity to photocopy it.” HN 6 “We make the comparison. […] If there are many zero-doses and under-vaccinated children, then we call the community health workers, we give them the tokens to go and collect […] the zero-doses and under-vaccinated children.” HN 7 “In the last six months, we’ve only held two monitoring meetings. The rest of the time, in other months, they weren’t available.” HN 1 “Usually, we develop the microplans ourselves within the health area. […] But this year, it wasn’t like that. The Health Zone Central Office developed the microplans for us to share. […] So far, I don’t even have the version […] The electronic version isn’t complete. It’s just pages. […] The recommendations are verbal. […] I forget. And the implementation is hypothetical. […] But if it’s written down […] I read what I was told […] and I start to understand what we were told.” HN 4 “If I finish, I make a photocopy. Sometimes […] I take photos from my phone. If I don’t have the money to photocopy them. So I have others on my phone, others I photocopy.” HN 1

### Subtheme 3: Functional staff availability and work organisation

The nominal number of providers did not always reflect the workforce actually available to deliver vaccination activities. On-call duties, rest periods, rotations, staff turnover and competing responsibilities reduced functional availability.

> “All five are private. The staff, either on rotation or if they’ve been on call, are off duty. […] Because in private facilities, staff changes frequently. Perhaps this person isn’t really suitable for vaccination. That also causes us problems.” HN 1 “In a private facility, there are certain obligations. You can’t just hire too many people. You’ll pay them for that. In the EPI, things are suffering. […] At the satellite clinics, if one of the people there isn’t present, the activity won’t take place. […] It’s suffering.” HN 3 “We might be told we have a monitoring meeting tomorrow, and someone will call to say they’ll be at the centre alone […] So, the time to sacrifice is impossible. It might be an overload problem […] They end up alone at the centre…” HN 6 “Staff shortage. […] She works alone. She’s the owner.” She’s the one who administers the vaccines. She’s the one who does everything. […] Sometimes, if community health workers aren’t available, she can’t leave the hospital. HN 7 “There’s a shortage of people, of service providers. In some facilities, there aren’t enough providers. So if you call her here, the person has to close the facility to come and meet you. […] Instead, I have to go there.” HN 4

### Subtheme 4: Operational resources, logistics and the cold chain

Continuity of vaccination depended on the practical availability of vaccines, transport, storage capacity and energy to maintain the cold chain. These conditions varied substantially across cases.

> “We have refrigerators, but they’re solar-powered. […] EPI-certified.” HN 3 “Our boss, the BDOM […] provided us with a substantial solar power kit. […] And we equipped the refrigerator itself with a solar kit. […] So, as far as the refrigerator is concerned, there are no issues with electricity.” HN 4 “If there aren’t any here, we make do on the ground. I ask the neighboring health centre, my colleagues, whoever has the vaccine can give it to me. I work, and the day I get supplies, I give it to them.” HN 6 “We coordinate with the facilities, we share the leftover vaccines from those days.” HN 3 “We don’t have refrigerators. The health centres are newly established. […] Not yet equipped. […] You were only told that you would organize the activities. […] Almost two years.” “HN 1 We had to get organized, we bought a refrigerator […] we also bought the panel and the converter. I raised the money at the health centre. […] We need to start storing a certain quantity of vaccines here.” HN 7

### Subtheme 5: Organisation of the community interface and adaptation of service delivery

Community health workers played a central role in identifying children, mobilising households and following up zero-dose and under-vaccinated children. Their contribution, however, depended on their availability and the support they received.

> “When they’re not paid for their activities, they work, and that impacts other activities and routines. Often, I only work with my president on routines.” HN 1 “We have some community health workers who have become discouraged in community activities related to vaccination. […] We were promised we would pay for it. But when it comes time to pay, they aren’t paid. […] That has discouraged the community health workers.” HN 4 “He goes out into the field […] two or three times a week to pick up children who have received no doses and those who are under-vaccinated.” HN 5 “If we make these comparisons, we find that there are many children who have received no doses and those who are under-vaccinated. So, we call the community health workers […] to pick up […] the children who have received no doses and those who are under-vaccinated.” HN 7

### Theme 3: Cultural factors

Two subthemes were identified: professional norms and variable uptake of EPI requirements; and perceptions, language and trust surrounding vaccination.

### Subtheme 6: Professional norms and variable uptake of EPI requirements

Participants described variation in the extent to which programme requirements were incorporated into professional practice, particularly in relation to reporting, implementation of recommendations and notification procedures.

> “Only serious cases are notified, but it’s recommended that they be notified.” HN 1 “There’s also negligence. […] He knows, he’s a former head nurse. […] It’s just negligence. I’ve already spoken with his medical director. Nothing changes. […] “To implement, you really have to be there every step of the way. […] The head nurses who participate in the Health Zone Central Office meetings, they find it easy to put the recommendations into action. […] Even if you tell them, to implement, you really have to talk, talk, talk.” HN 7

### Subtheme 7: Perceptions, language and trust surrounding vaccination

Participants also described social representations and communication barriers that could shape relationships between services and families.

> “We’ve politicized vaccination activities within the population. […] There aren’t any identified pockets of resistance. These are isolated cases… When you enter a household, everyone has their own reason. There are those who tell you that vaccines harm children’s health… You enter another household and they tell you that your vaccine will prevent our children from being born… Our children will be sterile.” HN 4 “The community health workers came to report a case of resistance and they even gave me the address. The families say they’re used to the ‘mishi’ roots; they don’t take vaccines. […] When we organize the campaigns, there has to be someone from Kasai, someone who speaks Tshiluba. At least they can manage to convince them, but for someone who speaks Swahili, it’s very difficult. […] Even here, during consultations, we have to bring in a Kasai nurse to interpret.” HN 7

### Theme 4: Exogenous factors

Two subthemes were identified: vertical dependence on coordination and decisions at health-zone level; and external dependence on routine resources, with transfer of costs to the periphery.

### Subtheme 8: Vertical dependence on coordination and decisions at health-zone level

Several local activities remained closely dependent on procedures, validation processes and decisions made by the Health Zone Central Office.

> “No, it’s at the Health Zone Central Office level. Because it’s the Health Zone Central Office that calls us. They’re developing the micro-plan. The people who will participate are the head nurse and the health-area committee chair.” HN 1 “He hasn’t decentralized the activity. […] Especially this time, he himself is the one who validates and serves. […] You leave like that, and he tells you he hasn’t filled out your document yet, you have to go back.” HN 3 “The central office authorized us; whoever doesn’t report back to us, we won’t give the vaccine anymore.” HN 7

### Subtheme 9: External dependence on routine resources and transfer of costs to the periphery

Financing represented another form of dependence. Participants distinguished between resources included in activity plans and funds actually available for implementation.

> “We plan for a certain amount of money for a given activity. […] But in fact, we don’t always have that money. […] And that remains the responsibility of the organisation if it has to be spent. […] The organisation also finances vaccination activities.” HN 4 “It’s the Health Zone Central Office that’s supposed to bring us […] But when it comes to EPI, we go down ourselves. We pay for our own transportation. […] We pick up. We bring back.” HN 3 “It’s been a whole year since we’ve had any forms. […] It’s a lack of support. To get the forms, we have to find ways to get them in other areas, here and there.” HN 2 “Yes, yes. But we weren’t given the money.” HN 7

### Cross-cutting theme: Compensatory local adaptations to contextual constraints

Across all four dimensions of Leichter’s framework, participants described local arrangements developed when routine mechanisms were insufficient. This cross-cutting pattern was present in all seven cases and included documentary, digital, organisational, logistical, financial and interorganisational adaptations.

> “If I finish, I make the photocopy. […] I take the photos from my phone. If I don’t have the money to photocopy them, then I have others on my phone, and I photocopy them.” HN 1 “We can even make a small card that we give to the mother; when the form arrives, we’ll fill it out.” HN 2 “We coordinate with the facilities, the vaccines for those days, we share what’s left.” HN 3 “If there isn’t one here, we make arrangements on the ground. I ask the neighboring health centre, my colleagues; whoever has the vaccine can give it to me. I work, and the day I get supplies, I give it to them.” HN 6 “I thought, okay, I’ll adapt. I called my friends. I said, we’ll meet on the phone. Open your phones, connect. […] Since […] it gave us good results, we’re going to expand it.” “HN 4 We had to get organized, we bought a refrigerator […] we also bought the panel and the converter. I raised the money at the health centre. […] We need to start storing a certain quantity of vaccines here.” HN 7

## Discussion

This study found that routine EPI service delivery across health areas in Mumbunda was shaped by interacting situational, structural, cultural and exogenous factors. These influences did not operate independently; rather, they combined differently across health areas, so that facilities working under the same programme guidelines could have markedly different capacities to deliver services.

Within Leichter’s framework, situational factors refer to circumstances and events that may temporarily alter the conditions under which a policy is implemented (7). In our study, these included population mobility, seasonality and time constraints. This finding is consistent with Sakas et al., who showed across three countries that immunisation performance depended partly on programmes’ capacity to adapt delivery strategies to local realities (8), and with Lawrence et al., who found in Malawi and Mozambique that apparently similar barriers operated differently according to local configurations of staffing, outreach, vaccine availability and service organisation (10). The importance of mobility in urban settings also aligns with the review by Belt et al., which showed that urban averages can conceal mobile and underserved populations (36). In Kinshasa, Mwamba et al. reported that 40% of surveyed children had received their most recent vaccination outside their health zone of residence (37). This is consistent with our finding that administrative boundaries alone may not adequately define the population actually served. Bangura et al. similarly identified migration, distance, inconvenient service hours, parental work constraints and language barriers as recurrent obstacles to childhood vaccination in sub-Saharan Africa (38). In this setting, access is therefore best understood as dynamic, reflecting the interaction of territory, time and the capacity of services to respond to families’ circumstances.

Structural factors represented the largest group of findings and encompassed peripheral governance, planning, information, staffing, logistics and the community interface. A common feature was the gap between resources or arrangements that formally existed and those that could actually be mobilised for vaccination. This distinction is consistent with LaFond et al.’s analysis of 12 districts in Ethiopia, Cameroon and Ghana, where districts that improved coverage were not necessarily those with more infrastructure, but those with stronger implementation strategies, management capacity and ability to adapt activities to local needs (11). Our findings suggest that similar variation may occur at an even finer scale, between health areas within the same health zone. Microplanning illustrates this gap particularly well. Mafigiri et al. showed in Uganda that the value of a microplan depended on how it was developed, understood and used by providers rather than on its mere existence (39). In Mumbunda, some microplans were developed without meaningful peripheral participation, were retained at the main facility or were produced at a higher level. Functional staff availability followed a similar logic: nominal staffing became insufficient when providers also had on-call duties, curative-care responsibilities, meetings or other competing tasks. Evidence on supportive supervision in low- and middle-income countries likewise indicates that effectiveness depends on problem-solving support and the practical availability of staff, rather than simply on the prescribed frequency of supervisory visits (40,41). Community health workers were also part of this functional capacity. Ogutu et al. showed in Nepal, Senegal and Zambia that their contribution to immunisation depended on organisation, motivation and trust (42). Our findings further suggest that their time carries an opportunity cost; treating them as continuously available volunteers risks overstating the programme’s actual community capacity. More broadly, the pattern observed in Mumbunda resembles Haq et al.’s systems analysis of the EPI in Pakistan, where staffing, demographic information, logistics, financing, vertical priorities and the sociopolitical environment jointly shaped programme performance (9).

Cultural factors extended beyond household beliefs to include professional norms within the EPI. This distinction is important because it avoids framing immunisation challenges solely as a problem of demand. National findings by Ishoso et al. showed that reasons for non-vaccination in the DRC differed between zero-dose and under-vaccinated children: the former were more strongly associated with motivations and perceptions, whereas the latter were more often linked to practical barriers (16). This heterogeneity is consistent with our finding of localised rather than uniform resistance. At provider level, selective reporting of adverse events following immunisation is also consistent with qualitative evidence from Ghana showing that reporting practices are influenced by knowledge, understanding of procedures and the organisational environment (43). Our findings add a relational dimension: trust was mediated by who conveyed the message. A community intermediary who spoke the household’s language, a nurse able to interpret or a socially recognised person could facilitate communication. This observation aligns with Ogutu et al.’s findings on the importance of trust in community health workers (42).

Exogenous factors showed how decisions and resources originating outside the health area shaped local functioning. This dependence was not inherently negative: the Health Zone Central Office provided supervision, guidance and technical support. It became constraining, however, when validation, procurement or planning depended heavily on a small number of actors, or when responsibilities were transferred without corresponding resources. These findings resonate with experience from the DRC. The Mashako Plan identified coordination, service delivery, vaccine availability and monitoring as critical areas (14). Our study shows how such system-level categories were experienced at the periphery through uneven dissemination of microplans, difficulties in supervision, dependence on particular supply channels and variation in available resources. The gap between budgeted activities and funds actually available also extends the findings of Le Gargasson et al., who identified constraints in budgetary processes, disbursement, staff motivation and dependence on external financing in the DRC (15). The interface with service users was another important organisational dimension. Kayembe-Ntumba et al. showed that service- and follow-up-related conditions were associated with vaccination dropout in the DRC (17). Our findings on waiting time, provider availability and reminders similarly indicate that access is not solely geographical. The present study therefore complements our earlier systemic analysis of EPI bottlenecks in Mumbunda (20) by showing the local contextual conditions in which those bottlenecks arise and why they may take different forms across health areas.

The adaptations described across the seven cases are consistent with the literature on everyday health-system resilience. Gilson et al. have shown how frontline actors sustain service functioning through problem solving, professional relationships and routine organisational adjustments (44,45), while Biddle et al. identify adaptive capacity as a central component of health-system resilience (46). Our findings nevertheless warrant caution. Borrowing vaccines, paying transport costs personally, using temporary documentation or purchasing equipment locally may sustain activities in the short term, but can also shift the burden of structural deficiencies to peripheral facilities and workers. Local resilience should therefore not be equated automatically with good system performance; it may also signal that costs and risks are being absorbed by frontline actors and facilities.

Several implications follow. First, microplanning should be genuinely participatory and conducted close to the point of service delivery, with regular updating of demographic information, population mobility, access constraints and community knowledge. Evidence from LaFond, Mafigiri and Sakas indicates that local capacity to adapt strategies is central to effective immunisation delivery (8,11,39). Second, human-resource planning should reflect functional rather than nominal availability. Training more than one staff member for essential EPI functions, avoiding concentration of key competencies in a single individual and organising supervision around problem solving may reduce organisational vulnerability (40,41). Third, routine immunisation financing should explicitly cover operational delivery costs, including transport, reproduction of tools, energy, cold-chain maintenance, supervision and community mobilisation. Vaughan et al. showed that delivery costs account for a substantial share of immunisation costs and need to be incorporated into financial planning (47). Fourth, the contribution of community health workers should be institutionalised and supported more predictably; recall, active tracing and cultural mediation cannot sustainably depend on unpaid goodwill alone (42). Finally, local adaptations should be differentiated between practices that may usefully be institutionalised such as digital communication, inter-area mutual support and peer learning and practices that primarily signal system weakness, including repeated out-of-pocket spending by staff, improvised tools and precarious storage arrangements. Strengthening the EPI should therefore support adaptive capacity without normalising peripheral underfunding (44–46).

This study has several limitations. It drew on seven head nurses from a single urban health zone and used sampling for analytical diversity rather than statistical representativeness; the findings therefore cannot be numerically generalised to Lubumbashi or the DRC as a whole. Head nurses are well positioned to describe the interface between the Health Zone Central Office, health facilities and communities, but their accounts do not directly represent the perspectives of parents, vaccinators, community health workers, health-zone managers or partners. Statements concerning health-zone decisions, professional behaviour or financial resources should therefore be interpreted as participants’ accounts. Numerical proportions, amounts and frequencies mentioned during interviews were not independently measured and should not be interpreted as population estimates. Finally, the limited data-collection period did not capture the full range of seasonal, financial or organisational variation in EPI delivery. Transferability therefore depends on the similarity of settings, particularly urban health zones characterised by multiple public and private providers, high population mobility, constrained operational resources and strong dependence between peripheral and higher system levels.

## Conclusion

Routine EPI service delivery in the Mumbunda Health Zone was shaped by the interaction of situational, structural, cultural and exogenous factors. Population mobility, seasonal and temporal constraints, variation in governance and microplanning, functional staff availability, logistical resources, community organisation, professional norms, perceptions of vaccination and dependence on decisions and resources outside the health area combined to produce different service-delivery conditions even within the same health zone.

A central finding was the gap between formal arrangements and what was operationally available: microplans that were not consistently disseminated, staff who were nominally present but not always functionally available, budgeted resources that were difficult to access, dependence on particular supply channels and routine costs sometimes absorbed by facilities or health workers. Local adaptations helped maintain service continuity, but they should not be treated as a sustainable substitute for adequately supported health-system functions.

Improving routine immunisation therefore requires more than increasing supplies or applying uniform strategies. Priority actions include participatory and regularly updated microplanning, reliable financing of routine operational costs, functional availability of more than one staff member able to deliver vaccination, stronger vaccine and cold-chain logistics and more predictable support for community health workers.

Further research should incorporate the perspectives of parents, community health workers, vaccinators, health-zone managers and partners, and compare multiple health zones to assess the transferability of these findings. Longitudinal studies could also examine how contextual factors and local adaptations evolve over time and identify which adaptations can be institutionalised without normalising underfunding or shifting system responsibilities onto frontline actors.

## Data availability statement

De-identified data supporting the findings are available from the corresponding author on reasonable request and subject to applicable institutional conditions. Full qualitative transcripts are not publicly available because of the risk of participant re-identification.

## Patient consent for publication

Not applicable.

## Author contributions

All authors contributed to the study design. CKD, AMN, ASS, HML and DCK contributed to study design, data management and project oversight. CKD, PPM, PCN, DMB and IKO contributed to formal analysis and drafting of the original manuscript. All authors critically reviewed the manuscript, approved the final version and accept accountability for the work. CKD is the guarantor.

The authors are non-native English speakers. ChatGPT (OpenAI) was used solely for English-language editing to improve grammar, syntax, clarity and consistency. All AI-assisted edits were reviewed by the authors, who retained full responsibility for the accuracy, interpretation and conclusions of the manuscript.

## Patient and public involvement

Patients and/or the public were not involved in the design, conduct, reporting or dissemination plans of this research.

## Acknowledgements

We would like to thank the Senior Nurses in the health zones who agreed to take part in this study. We would also like to thank the Senior Nurses in the Kilimasimba 1 and Somika health zones, within the Kisanga Health Zone, for providing us with the opportunity to pre-test the data collection tool.

## REFERENCES

1. Petu A, Masresha B, Wiysonge CS, Mwenda J, Nyarko K, Bwaka A, et al. Reflections on 50 years of immunisation programmes in the WHO African region: an impetus to build on the progress and address the unfinished immunisation business. BMJ Glob Health. 21 mai 2025;10(5). doi:10.1136/bmjgh-2024-017982 PubMed PMID: 10.1136/bmjgh-2024-017982.

2. World Health Organization. Challenges in global immunization and the Global Immunization Vision and Strategy 2006-2015= Les enjeux de la vaccination dans le monde: perspectives et stratégies 2006-2015. Wkly epidemiol rec. 2006;190⍰5.

3. Shattock AJ, Johnson HC, Sim SY, Carter A, Lambach P, Hutubessy RCW, et al. Contribution of vaccination to improved survival and health: modelling 50 years of the Expanded Programme on Immunization. The Lancet. 25 mai 2024;403(10441):2307⍰16. doi:10.1016/S0140-6736(24)00850-X PubMed PMID: 38705159.

4. Gilson L, Alliance pour la recherche sur les politiques et systèmes de santé, OMS. Recherche sur les politiques et systèmes de santé : manuel de méthodologie□: version abrégée. Service de production des documents de l’OMS. Genève, Suisse: OMS; 2013. 112 p.

5. Gilson L, Hanson K, Sheikh K, Agyepong IA, Ssengooba F, Bennett S. Building the Field of Health Policy and Systems Research: Social Science Matters. PLOS Medicine. 23 août 2011;8(8):e1001079. doi:10.1371/journal.pmed.1001079

6. Papanicolas I, Rajan D, Karanikolos M, Soucat A, Figueras J, éditeurs. Health system performance assessment: A framework for policy analysis [Internet]. Copenhagen (Denmark): European Observatory on Health Systems and Policies; 2022 [cité 22 juill 2025]. (European Observatory Health Policy Series). Disponible sur: http://www.ncbi.nlm.nih.gov/books/NBK590192/ PubMed PMID: 37023239.

7. Leichter HM. A Comparative Approach to Policy Analysis: Health Care Policy in Four Nations [Internet]. 1979 [cité 26 juill 2025]. Disponible sur: https://repository.library.georgetown.edu/handle/10822/782133

8. Sakas Z, Hester KA, Ellis A, Ogutu EA, Rodriguez K, Bednarczyk R, et al. Critical success factors for high routine immunisation performance: a qualitative analysis of interviews and focus groups from Nepal, Senegal, and Zambia. BMJ Open. 4 oct 2023;13(10):e070541. doi:10.1136/bmjopen-2022-070541 PubMed PMID: 37793916; PubMed Central PMCID: PMC10551940.

9. Haq Z, Shaikh BT, Tran N, Hafeez A, Ghaffar A. System within systems: challenges and opportunities for the Expanded Programme on Immunisation in Pakistan. Health Res Policy Syst. 17 mai 2019;17(1):51. doi:10.1186/s12961-019-0452-z PubMed PMID: 31101060; PubMed Central PMCID: PMC6525435.

10. Lawrence E, Metje A, Matemba C, Powelson J. Identifying context-specific drivers of routine childhood immunisation dropout in Mozambique and Malawi: a secondary thematic analysis of qualitative community-based participatory research data. BMJ Open. 1 nov 2025;15(11):e104490. doi:10.1136/bmjopen-2025-104490 PubMed PMID: 41263846.

11. LaFond A, Kanagat N, Steinglass R, Fields R, Sequeira J, Mookherji S. Drivers of routine immunization coverage improvement in Africa: findings from district-level case studies. Health Policy Plan. 1 avr 2015;30(3):298⍰308. doi:10.1093/heapol/czu011

12. Adamu AA, Jalo RI, Masresha BG, Ndwandwe D, Wiysonge CS. Mapping the Implementation Determinants of Second Dose Measles Vaccination in the World Health Organization African Region: A Rapid Review. Vaccines (Basel). 8 août 2024;12(8):896. doi:10.3390/vaccines12080896 PubMed PMID: 39204023; PubMed Central PMCID: PMC11359529.

13. DRC-National Institute of Statistics, Kinshasa School of Public Health, ICF. Democratic Republic of the Congo Demographic and Health Survey 2023–2024: Final Report [Enquête Démographique et de Santé EDS-RDC III 2023–2024 : Rapport final] [Internet]. Kinshasa, RDC et Rockville, Maryland, USA : ICF.; 2025 [cité 18 août 2025]. Disponible sur: https://www.dhsprogram.com/pubs/pdf/SR291/SR291.pdf

14. Lame P, Milabyo A, Tangney S, Mbaka GO, Luhata C, Le Gargasson JB, et al. A successful national and multipartner approach to increase immunization coverage: the democratic republic of Congo Mashako plan 2018–2020. Global Health: Science and Practice [Internet]. 2023 [cité 24 mai 2025];11(2). Disponible sur: https://www.ghspjournal.org/content/11/2/e2200326.abstract

15. Le Gargasson JB, Mibulumukini B, Gessner BD, Colombini A. Budget process bottlenecks for immunization financing in the Democratic Republic of the Congo (DRC). Vaccine. 2014;32(9):1036 42.

16. Ishoso DK, Mafuta E, Danovaro-Holliday MC, Ngandu C, Menning L, Cikomola AMW, et al. Reasons for Being “Zero-Dose and Under-Vaccinated” among Children Aged 12–23 Months in the Democratic Republic of the Congo. Vaccines. août 2023;11(8):8. doi:10.3390/vaccines11081370

17. Kayembe-Ntumba HC, Vangola F, Ansobi P, Kapour G, Bokabo E, Mandja BA, et al. Vaccination dropout rates among children aged 12-23 months in Democratic Republic of the Congo: a cross-sectional study. Arch Public Health. 5 janv 2022;80(1):18. doi:10.1186/s13690-021-00782-2

18. Acharya P, Kismul H, Mapatano MA, Hatløy A. Individual- and community-level determinants of child immunization in the Democratic Republic of Congo: A multilevel analysis. PLoS One. 2018;13(8):e0202742. doi:10.1371/journal.pone.0202742 PubMed PMID: 30138459; PubMed Central PMCID: PMC6107214.

19. Bureau Central de la Zone de Santé de Mumbunda. Micro plan des activités de vaccination PEV de routine de la Zone de Santé de Mumbunda 2026. 2026.

20. Diyoka CK, Kalombola DC, Saidi AS, Mukanga PP, Omba IK, Nkola AM. Systemic analysis of bottlenecks in routine EPI coverage vaccination among children aged 0–23 months in the health areas of the Mumbunda Health District: a cross-sectional mixed-methods study [Internet]. Research Square; 2026 [cité 12 sept 2026]. Disponible sur: https://www.researchsquare.com/article/rs-10651748/v1 doi:10.21203/rs.3.rs-10651748/v1

21. Sheikh K, Gilson L, Agyepong IA, Hanson K, Ssengooba F, Bennett S. Building the Field of Health Policy and Systems Research: Framing the Questions. PLOS Medicine. 16 août 2011;8(8):e1001073. doi:10.1371/journal.pmed.1001073

22. Ecole de santé publique-Unilu, Ministere de la santé, Institut of Tropical Medicine Antwerp, UrbanBirth Collective. figshare [figure] [Internet]. figshare; 2025 [cité 24 oct 2025]. Detailed maps of health facilities, health zones and health areas in Lubumbashi city, 2023. Disponible sur: https://figshare.com/s/87e6f3140a88340e0663

23. Ministère de la Santé. Recueil des normes de création, d’organisation et de fonctionnement des structures de la zone de santé en République démocratique du Congo [Internet]. Secrétariat général, Ministere de la santé publique; 2019 [cité 11 oct 2025]. Disponible sur: https://bv-assk.org/wp-content/uploads/2024/03/Recueil-des-normes-de-creation-dorganisation-de-fonctionnement-des-structures-de-la-ZS-en-RDC-MSP-2019.pdf

24. Ministère de la Santé Publique-RDC. Stratégie de Renforcement du système de santé. Kinshasa; 2006. p. 1⍰50. Rapport Deuxieme édition.

25. Dunia GMB. Implantation des sites de soins communautaires en R□publique D□mocratique du Congo: cons□cration d□un double standard dans l□acc□s aux soins. PAMJ Clinical Medicine. 24 avr 2013;14(158). doi:10.11604/pamj.2013.14.158.2003

26. Chenge M, Van der Vennet J, Porignon D, Luboya N, Kabyla I, Criel B. La carte sanitaire de la ville de Lubumbashi, République Démocratique du Congo Partie I□: problématique de la couverture sanitaire en milieu urbain congolais. Glob Health Promot. 1 sept 2010;17(3):63⍰74. doi:10.1177/1757975910375173

27. Sambo L, Chatora R, Goosen E. Tools for assessing the operationality of district health systems. Brazzaville: World Health Organization, Regional Office for Africa [Internet]. 2003 [cité 31 oct 2025]. Disponible sur: https://indexmedicus.afro.who.int/iah/fulltext/health%20systems1.pdf

28. Durán A, Kutzin J, Martin-Moreno JM, Travis P. Understanding health systems: scope, functions and objectives. Health systems: Health, wealth, society and wellbeing Maidenhead, Open University Press and McGraw-Hill. 2011;19⍰36.

29. OMS. Rapport sur la santé dans le monde, 2000: Pour un système de santé plus performant [Internet]. Geneve: Organisation mondiale de la Santé; 2000 [cité 4 juill 2025]. Disponible sur: https://apps.who.int/gb/archive/pdf_files/wha53/fa4.pdf

30. Van Olmen J, Criel B, Van Damme W, Marchal B, Van Belle S, Van Dormael M, et al. Analysing health systems to make them stronger. Studies in health services organisation and policy-Antwerp, 1997, currens [Internet]. 2010 [cité 3 juill 2025]. Disponible sur: https://repository.uantwerpen.be/docman/irua/2643c6/159213.pdf

31. Bandara S, Phiri MM, Magati P, Drope J, Adams A, Hunt M, et al. Contextual factors impacting WHO Framework Convention on Tobacco Control implementation in Africa—a scoping review. Health Promot Int. 1 déc 2024;39(6):daae155. doi:10.1093/heapro/daae155

32. StraGlaser G, Strauss AL. The discovery of grounded theory: Strategies for qualitative research. New Bruinswick and Lond. Routledge; 1967.

33. Naeem M, Ozuem W, Howell K, Ranfagni S. Demystification and Actualisation of Data Saturation in Qualitative Research Through Thematic Analysis. International Journal of Qualitative Methods. 1 mai 2024;23:16094069241229777. doi:10.1177/16094069241229777

34. Braun V, Clarke V. Using thematic analysis in psychology. Qualitative Research in Psychology. 1 janv 2006;3(2):77⍰101. doi:10.1191/1478088706qp063oa

35. Braun V, Clarke V. One size fits all? What counts as quality practice in (reflexive) thematic analysis? Qualitative Research in Psychology. 3 juill 2021;18(3):328⍰52. doi:10.1080/14780887.2020.1769238

36. Belt RV, Abdullah S, Mounier-Jack S, Sodha SV, Danielson N, Dadari I, et al. Improving Equity in Urban Immunization in Low- and Middle-Income Countries: A Qualitative Document Review. Vaccines. juill 2023;11(7):1200. doi:10.3390/vaccines11071200

37. Mwamba GN, Yoloyolo N, Masembe Y, Nsambu MN, Nzuzi C, Tshekoya P, et al. Vaccination coverage and factors influencing routine vaccination status in 12 high risk health zones in the Province of Kinshasa City, Democratic Republic of Congo (DRC), 2015. Pan Afr Med J. 21 juin 2017;27(Suppl 3):7. doi:10.11604/pamj.supp.2017.27.3.11930 PubMed PMID: 29296142; PubMed Central PMCID: PMC5745950.

38. Bangura JB, Xiao S, Qiu D, Ouyang F, Chen L. Barriers to childhood immunization in sub-Saharan Africa: A systematic review. BMC Public Health. 14 juill 2020;20(1):1108. doi:10.1186/s12889-020-09169-4

39. Mafigiri DK, Iradukunda C, Atumanya C, Odie M, Mancuso A, Tran N, et al. A qualitative study of the development and utilization of health facility-based immunization microplans in Uganda. Health Res Policy Sys. 11 août 2021;19(2):52. doi:10.1186/s12961-021-00708-y

40. Deussom R, Mwarey D, Bayu M, Abdullah SS, Marcus R. Systematic review of performance-enhancing health worker supervision approaches in low- and middle-income countries. Hum Resour Health. 6 janv 2022;20(1):2. doi:10.1186/s12960-021-00692-y

41. Rowe SY, Ross-Degnan D, Peters DH, Holloway KA, Rowe AK. The effectiveness of supervision strategies to improve health care provider practices in low- and middle-income countries: secondary analysis of a systematic review. Hum Resour Health. 6 janv 2022;20(1):1. doi:10.1186/s12960-021-00683-z

42. Ogutu EA, Ellis AS, Hester KA, Rodriguez K, Sakas Z, Jaishwal C, et al. Success in vaccination programming through community health workers: a qualitative analysis of interviews and focus group discussions from Nepal, Senegal and Zambia. BMJ Open. 1 avr 2024;14(4):e079358. doi:10.1136/bmjopen-2023-079358 PubMed PMID: 38569679.

43. Aborigo RA, Welaga P, Oduro A, Shaum A, Opare J, Dodoo A, et al. Optimising reporting of adverse events following immunisation by healthcare workers in Ghana: A qualitative study in four regions. PLoS One. 2022;17(12):e0277197. doi:10.1371/journal.pone.0277197 PubMed PMID: 36538549; PubMed Central PMCID: PMC9767370.

44. Gilson L, Ellokor S, Lehmann U, Brady L. Organizational change and everyday health system resilience: Lessons from Cape Town, South Africa. Soc Sci Med. déc 2020;266:113407. doi:10.1016/j.socscimed.2020.113407 PubMed PMID: 33068870; PubMed Central PMCID: PMC7538378.

45. Gilson L, Barasa E, Nxumalo N, Cleary S, Goudge J, Molyneux S, et al. Everyday resilience in district health systems: emerging insights from the front lines in Kenya and South Africa. BMJ Glob Health. 2017;2(2):e000224. doi:10.1136/bmjgh-2016-000224 PubMed PMID: 29081995; PubMed Central PMCID: PMC5656138.

46. Biddle L, Wahedi K, Bozorgmehr K. Health system resilience: a literature review of empirical research. Health Policy Plan. 1 oct 2020;35(8):1084⍰109. doi:10.1093/heapol/czaa032 PubMed PMID: 32529253; PubMed Central PMCID: PMC7553761.

47. Vaughan K, Ozaltin A, Mallow M, Moi F, Wilkason C, Stone J, et al. The costs of delivering vaccines in low- and middle-income countries: Findings from a systematic review. Vaccine X. 9 août 2019;2:100034. doi:10.1016/j.jvacx.2019.100034 PubMed PMID: 31428741; PubMed Central PMCID: PMC6697256.

